# The contribution of asymptomatic SARS-CoV-2 infections to transmission - a model-based analysis of the Diamond Princess outbreak

**DOI:** 10.1101/2020.05.07.20093849

**Authors:** Jon C Emery, Timothy W Russell, Yang Liu, Joel Hellewell, Carl AB Pearson, CMMID 2019-nCoV working group, Gwenan M Knight, Rosalind M Eggo, Adam J Kucharski, Sebastian Funk, Stefan Flasche, Rein M G J Houben

**Author notes:** The following authors were part of the Centre for Mathematical Modelling of Infectious Disease 2019-nCoV working group. Each contributed in processing, cleaning and interpretation of data, interpreted findings, contributed to the manuscript, and approved the work for publication: Katherine E. Atkins, Petra Klepac, Akira Endo, Christopher I Jarvis, Nicholas G. Davies, Eleanor M Rees, Sophie R Meakin, Alicia Rosello, Kevin van Zandvoort, James D Munday, W John Edmunds, Thibaut Jombart, Megan Auzenbergs, Emily S Nightingale, Mark Jit, Sam Abbott, David Simons, Nikos I Bosse, Quentin J Leclerc, Simon R Procter, C Julian Villabona-Arenas, Damien C Tully, Arminder K Deol, Fiona Yueqian Sun, Stéphane Hué, Anna M Foss, Kiesha Prem, Graham Medley, Amy Gimma, Rachel Lowe, Samuel Clifford, Matthew Quaife, Charlie Diamond, Hamish P Gibbs, Billy J Quilty, Kathleen O’Reilly*. Note: paper <u>not yet</u> peer reviewed. Corresponding author: Rein Houben.

## Abstract

**Background:** Some key gaps in the understanding of SARS-CoV-2 infection remain. One of them is the contribution to transmission from individuals experiencing asymptomatic infections. We aimed to characterise the proportion and infectiousness of asymptomatic infections using data from the outbreak on the Diamond Princess cruise ship.

**Methods:** We used a transmission model of COVID-19 with asymptomatic and presymptomatic states calibrated to outbreak data from the Diamond Princess, to quantify the contribution of asymptomatic infections to transmission. Data available included the date of symptom onset for symptomatic disease for passengers and crew, the number of symptom agnostic tests done each day, and date of positive test for asymptomatic and presymptomatic individuals.

**Findings:** On the Diamond Princess 74% (70-78%) of infections proceeded asymptomatically, i.e. a 1:3.8 case-to-infection ratio. Despite the intense testing 53%, (51-56%) of infections remained undetected, most of them asymptomatic. Asymptomatic individuals were the source for 69% (20-85%) of all infections. While the data did not allow identification of the infectiousness of asymptomatic infections, assuming no or low infectiousness resulted in posterior estimates for the net reproduction number of an individual progressing through presymptomatic and symptomatic stages in excess of 15.

**Interpretation:** Asymptomatic SARS-CoV-2 infections may contribute substantially to transmission. This is essential to consider for countries when assessing the potential effectiveness of ongoing control measures to contain COVID-19.

**Funding:** ERC Starting Grant (#757699), Wellcome trust (208812/Z/17/Z), HDR UK (MR/S003975/1)

## Research in context

### Evidence before this study

It is known that a non-trivial proportion of infections with SARS-CoV-2 remain asymptomatic, and there is evidence that asymptomatic individuals contribute to transmission. However, empirical estimates for the proportion of infections that remain asymptomatic are often difficult to interpret due to opportunistic sampling frames combined with low and imbalanced participation from individuals with and without symptoms, which have resulted in a wide range of values (between 6-96%), with a suggestion of variation across age-groups. Quantitative estimates for the contribution of asymptomatic SARS-CoV-2 infections to ongoing transmission are absent.

### Added value of this study

In this study we calibrated a mechanistic transmission model to data from the Diamond Princess cruise ship outbreak, which is unique in that it occurred in a closed population, nearly all individuals were tested regardless of symptoms at least once and detailed open access data are available. Our data-driven model found that 74% (95% posterior interval (PI) = 70-78%) of SARS-CoV-2 infections proceeded asymptomatically, a case-infection ratio of 1:3.8 (1:3.3-1:4.4). We found that because systematic testing irrespective of symptoms was only implemented in the last days before disembarkation, over half (53%, (51-56%) of infected individuals were not detected during this outbreak.

Our model provides the first quantitative estimates of the proportion of all transmission driven by asymptomatic individuals. In a context of rapid and near complete case-isolation as well as quarantine, asymptomatic infections cases were responsible for 69% (20-85%) of all new infections. Remaining transmission was equally distributed between the presymptomatic and symptomatic phases of COVID-19 which is in line with previous findings. Part of the remaining uncertainty is due to the relative infectiousness of asymptomatic individuals, which we were unable to estimate. However, an exploration of the scenarios with a low relative infectiousness (e.g. 0-25% compared to symptomatic individuals) showed that to replicate the data a very high net reproduction number was required for individuals progressing through presymptomatic and symptomatic stages (15.5-29.1).

### Implications of all the available evidence

In this outbreak, the majority of infections proceeded asymptomatically, and remained mostly undetected. Asymptomatic individuals likely contributed substantially to SARS-CoV-2 transmission. Hence, control measures, and models projecting their potential impact, need to look beyond the symptomatic cases if they are to understand and address ongoing transmission.

## Introduction

The ongoing COVID-19 pandemic has spread rapidly across the globe, and the number of individuals infected with SARS-CoV-2, outstrips the number of reported cases (1,2). One key reason for this may be that a substantial proportion of cases proceed asymptomatically, i.e. they either do not experience, or are not aware of symptoms throughout their infection but despite that can transmit to others. In this sense, asymptomatic infections differ from presymptomatic ones, which describes the part of the incubation period before symptoms develop during which onward transmission is possible.

While pre- and asymptomatic individuals do not directly contribute to morbidity or mortality in an outbreak, they can contribute to ongoing transmission, as has been shown for COVID-19, (3–5) and other diseases (6–8). Particularly, purely symptom-based interventions (e.g., self-isolation upon onset of disease) will not interrupt transmission from asymptomatic individuals and hence may be insufficient for outbreak control if a substantial proportion of transmission originates from pre- and asymptomatic infections. (4)

An estimate of the proportion of infections that never progress to symptomatic disease, also known as the case-to-infection ratio, provides an indicator of what proportion of cases will remain undetected by symptom-based case detection (9). Evidence so far has suggested that the proportion of SARS-CoV-2 infections that proceed asymptomatically is likely non-trivial (10–15), although empirical data is often difficult to interpret due to opportunistic sampling frames (16) combined with low (17) and imbalanced participation from individuals with and without symptoms (18). While it is likely that transmission from asymptomatic individuals can occur, (19) quantitative estimates are effectively absent. Improved understanding of the relative infectiousness of asymptomatic SARS-CoV-2 infection, and its contribution to overall transmission will greatly improve the ability to estimate the impact of intervention strategies. (9) What is known is that in the presence of active case-finding, presymptomatic infections and symptomatic cases contribute almost equally to overall transmission, as both modelling and empirical studies have shown. (11,12)

Documented outbreaks in a closed population with extensive testing of individuals regardless of symptoms provide unique opportunities for improved insights into the dynamics of an infection, as knowledge of the denominator and true proportion infected are crucial, yet often unavailable in other datasets. Here, we use data from the well-documented outbreak on the Diamond Princess cruise ship to capture the mechanics of COVID-19 in a transmission model to infer estimates for the proportion, infectiousness and contribution to transmission of asymptomatic infections.

## Methods

### Data

Data from the Diamond Princess outbreak have been widely reported (13,20,21). On January 20th, the Diamond Princess cruise ship departed from Yokohama on a tour of Southeast Asia. A passenger that disembarked on January 25th in Hong Kong subsequently tested positive for SARS-CoV-2 on February 1st, reporting the date of symptom onset as January 23rd.

After arriving back in Yokohama on February 3rd, all passengers and crew were screened for symptoms, and those screening positive were then tested. The ship began quarantine on February 5th with all passengers confined to their cabins and crew undertaking essential activities only. At the start of quarantine there were 3,711 individuals on board (2,666 passengers and 1,045 crew) with a median age of 65 (45-75 interquartile range).

Testing capacity was limited until February 11th and before then the majority of individuals tested had reported symptoms, referred to here as ‘symptom-based testing’. All individuals with a positive test at any stage were promptly removed from the ship and isolated. After February 11th, testing capacity increased and the testing of individuals irrespective of symptoms, referred to here as ‘symptom-agnostic testing’, was scaled up. In total, 314 symptomatic and 320 pre- or asymptomatic infections were reported before disembarkation was principally completed on February 21st.

We extracted the following data from (13,20,21) (**see Figure 1**). Firstly, the number of symptomatic cases per day (i.e. those testing positive having reported symptoms) by date of symptom onset, separately for passengers and crew. The date of symptom onset was not available for 115 cases, which we accounted for in our model structure by assuming they were distributed over time proportional to those cases with a reported date of symptom onset (see supplementary materials for details). Secondly, we extracted the number of pre- or asymptomatic infections identified per day (i.e. individuals testing positive having not reported symptoms) by date of test. The test date was not available for 35 pre- or asymptomatic individuals between the February 6-14th, which we assumed were distributed over time proportional to the daily number of tests performed amongst individuals not reporting symptoms. No data were available on how many individuals that tested positive in the absence of symptoms became symptomatic after disembarkation. Finally, we extracted the number of tests performed per day amongst individuals not reporting symptoms.

**Figure 1:**
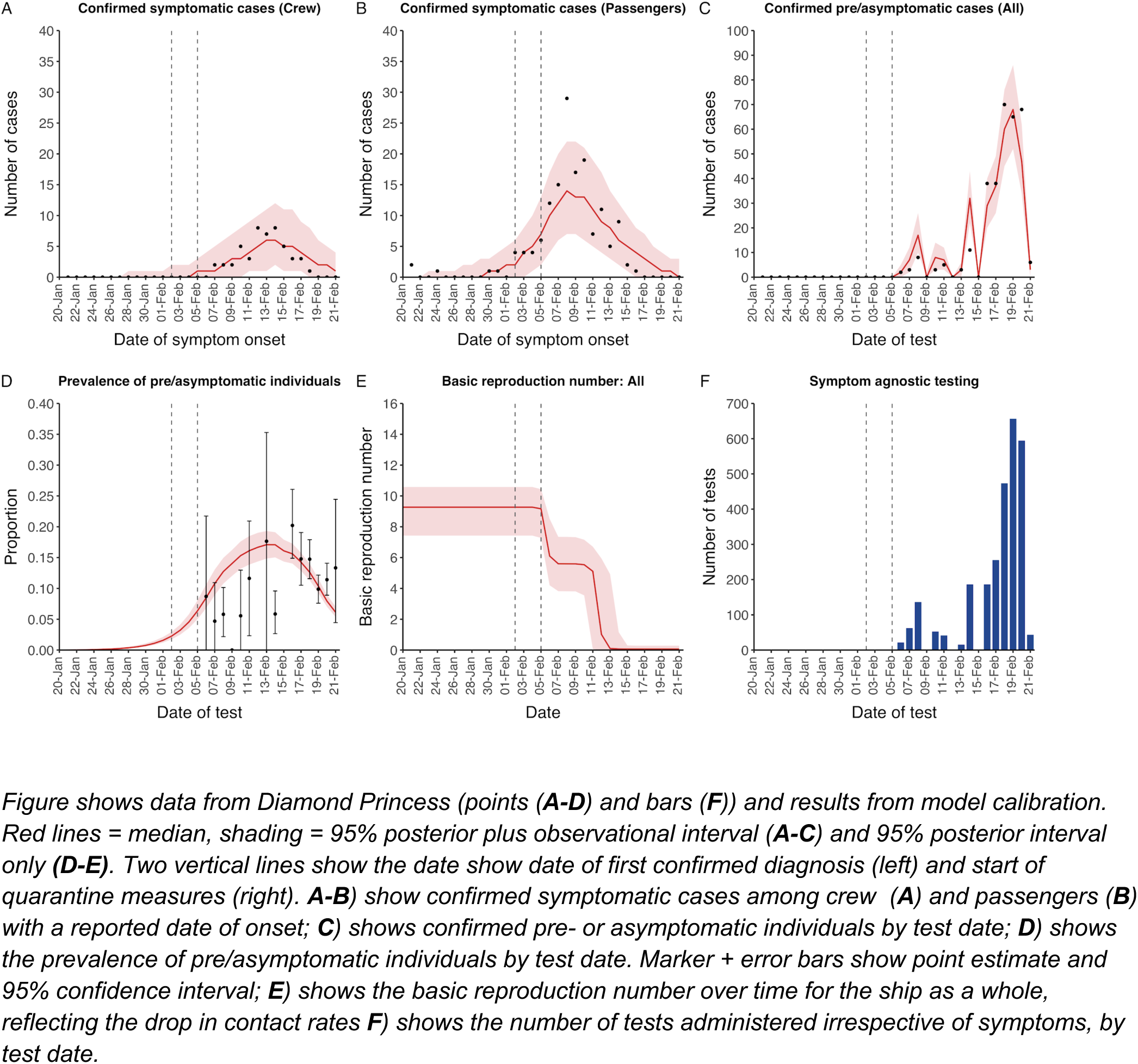
Data from Diamond Princess and model calibration.

### Model

We built a deterministic, compartmental model to capture transmission, disease development and the effect of interventions on board the Diamond Princess. Following exposure, after which an individual is assumed to test negative for SARS-CoV-2 for the duration of the latent phase (see **table 1**), a proportion of individuals proceed asymptomatically with the remainder becoming presymptomatic. Individuals in the presymptomatic, asymptomatic or symptomatic state are assumed to test positive and have independent infectiousness, expressed relative to those with symptomatic disease.

**Table 1:** Model parameters and priors/values.

| Parameter | Description | Prior/value | Source/Notes |
| --- | --- | --- | --- |
| $\bar{\beta}$ | Overall contact rate (1/days) | Estimated: *Uniform*(0,100) | |
| $c^{(cc)}$ | Relative initial contact rate between crew/crew | Fixed: 1 | |
| $c^{(pp)}$ | Relative initial contact rate between passengers/passengers | Estimated: *Uniform*(0,100) | |
| $c^{(pc)}$ | Relative initial contact rate between passengers/crew | Fixed relative to $c^{(pp)}$ | |
| $X$ | Ratio: $\frac{c^{(pc)}}{c^{(pp)}}$ | Fixed: 0.1 | Assumed. Varied in sensitivity analyses |
| $b_1$ | Percentage reduction in all initial contact rates (%) | Estimated: *Uniform*(0,100) | |
| $b_2$ | Rate of change of all contact rates (1/days) | Fixed: 10 | Assumed. Transitions completed over approximately one day |
| $\tau^{(pp)}, \tau^{(pc)}$ | Time of transition for contacts between passengers/passengers and passengers/crew (days) | Estimated: *Uniform*(0,32) | Assumed to be equal to each other |
| $\tau^{(cc)}$ | Time of transition for contacts between crew/crew (days) | Estimated: *Uniform*(0,32) | |
| $\theta_p$ | Relative infectiousness of presymptomatic state | Estimated: *Uniform*(0,1) | Relative to symptomatic state |
| $\theta_a$ | Relative infectiousness of asymptomatic state | Estimated: *Uniform*(0,1) | Relative to symptomatic state |
| $\chi$ | Proportion of infections that proceed to asymptomatic state | Estimated: *Uniform*(0,1) | |
| $\frac{1}{\nu}$ | Latent period (days) | Fixed: 4.3 | Derived from (23) |
| $\frac{1}{\gamma_a}$ | Mean duration in asymptomatic state (days) | Fixed: 5.0 | Assumed. Sum of mean durations in presymptomatic and symptomatic states. Varied in sensitivity analyses. |
| $\frac{1}{\gamma_D}$ | Mean duration in presymptomatic state (days) | Fixed: 2.1 | Derived from (23) |
| $\frac{1}{\gamma_s}$ | Mean duration in infectious symptomatic state (days). | Fixed: 2.9 | From (24) Applicable only until quarantine starts on 5th Feb |
| $\frac{1}{\mu}$ | Mean delay between onset of symptomatic disease and symptom based-testing and removal (days). | Fixed: 1 | Assumed. Applicable only after quarantine starts on 5th Feb. |
| $\frac{1}{\eta}$ | Mean duration of test positivity following recovery (days) | Fixed: 7 | From (22) |
| $\phi$ | Proportion of symptomatic cases with a reported onset date | Fixed: 0.661 (199/314) | From (13,21) |
| $f(t)$ | Rate of symptom-agnostic testing and removal (1/days) | Fixed: *Calculated* | From (13) Calculated using the number of tests administered per day amongst individuals not reporting symptoms (see **Supplementary Materials**) |
| $N^{(p)}$ | Total number of passengers on the ship as at start of quarantine on 5th Feb | Fixed: 2,666 | From (20) |
| $N^{(c)}$ | Total number of crew on the ship as at start of quarantine on 5th Feb | Fixed: 1,045 | From (20) |

Individuals with presymptomatic infection are either detected through symptom-agnostic testing before being removed from the ship, or develop symptomatic disease. Once symptomatic disease starts, individuals can either recover undetected on the ship or, following the start of quarantine on February 5th, be detected through symptom-based testing and removed from the ship with an average delay of one day following symptom onset. We allowed for individuals to test positive after their infectious period for an average of seven days. (22) After this, we assume they would test negative.

Individuals with asymptomatic infections either recover undetected on the ship, or are detected by symptom-agnostic testing before being removed from the ship. See supplementary figure 1 for model diagramme. The proportion of pre- and asymptomatic individuals tested and removed from the ship per day was determined by the number of tests performed per day amongst individuals not reporting symptoms (**Figure 1F**).

Crew and passengers were modelled separately, using stratified data on the number of confirmed symptomatic cases (**Figure 1A-B**). We estimated the within-crew and within-passenger contact rates through calibration to the data, but assumed that the between group contact rate was a fixed factor of 1/10th of the within-group rate, and explored the impact of this assumption in sensitivity analyses (see supplementary materials). We enabled the model to capture potential changes in contact behaviour between individuals by representing contact rates as sigmoid functions over time, reflecting reductions in contact. The dates and extent of the changes were determined solely through model calibration to the data.

### Model parameterisation

We used data from the literature to inform the natural history of COVID-19, in particular for the duration of presymptomatic and symptomatic phases (Table 1).

### Model calibration

The model was calibrated in a Bayesian framework. We fitted to the daily incidence of confirmed symptomatic cases with a known onset date, separately for passengers and crew, assuming a Poisson distribution in the likelihood. We simultaneously fitted to the daily number of confirmed pre- and asymptomatic infections for passengers and crew combined by using the number of tests administered per day and the prevalence of presymptomatic, asymptomatic and post-infection test-positive individuals, assuming a binomial distribution in the likelihood. We used uniform priors for the parameters to be estimated (see Table 1) and sampled the posterior of the model parameters using sequential Markov Chain Monte-Carlo (MCMC). A burn in phase during which the proposal distributions were adapted in both scale and shape to provide optimal sampling efficiency was discarded, leaving chains with one million iterations. The resultant MCMC chains were visually inspected for convergence (see supplementary materials for details).

### Model outputs

Model outputs were calculated by randomly sampling 100,000 parameter values from the posterior distribution. Model trajectories were generated and compared to the data in **Figure 1A-C** to inspect model fit. The basic reproduction number was also calculated over time, as a measure of ongoing transmission. We estimated the proportion of infections that become asymptomatic and the relative infectiousness of asymptomatic infections using their respective marginal posterior parameter values. Finally, the contribution of asymptomatic infections to overall transmission, as well as the net reproduction number for presymptomatic passengers at the beginning of the outbreak (i.e. the typical number of infections generated by a single presymptomatic individual) were estimated, both overall and by specific ranges of relative infectiousness. We report the median and 95% equal-tailed posterior intervals (PI) throughout.

### Sensitivity analyses

We recalibrated the model for a number of alternative scenarios to assess model sensitivity. Firstly, we assessed the impact of removing the asymptomatic phase (i.e. 100% of infections progressed to symptomatic disease). Secondly, we explored the impact of assuming different values for the relative mixing between crew and passengers as well as shorter and longer durations of asymptomatic infection. Thirdly, we explored the impact of assuming a different proportion of asymptomatic infections for crew and passengers based on their distinct median ages (36 years for crew, 69 years for passengers), using a fixed ratio for the two proportions taken from the results of a model fitted to epidemic data in six countries by Davies et al. (25) In addition, we explored a longer latent period given the relatively high age in our population. Finally, we recalibrated the model assuming the 35 confirmed pre/asymptomatic cases where a test date was not available were allocated to the last feasible day (13th Feb) instead of proportionate to the overall number of tests over the period February 6-14th. See supplementary materials for further details. (26) See supplementary materials for further details.

All analyses were conducted using R version 3.5.0 (27). Bayesian calibration was performed in LibBi (28) using RBi (29) as an interface. Replication data and analyses scripts are available on GitHub at https://github.com/thimotei/covid19_asymptomatic_trans.

### Role of funding source

The funder of the study had no role in study design, data collection, data analysis, data interpretation, or writing of the report. The corresponding author had full access to all the data in the study and had final responsibility for the decision to submit for publication.

## Results

### Calibration

The model reflected the data well (**Figure 1**), including the differently timed peaks for confirmed symptomatic cases for passengers (**Figure 1A**) and crew (**Figure 1B**). In addition, the model matched the expected impact of quarantine of passengers on transmission from February 4th as illustrated by the drop in reproductive number, **Figure 1E**), followed by a later drop in transmission after February 10th, which was driven by a change in contact pattern in crew. See **supplementary materials** for full MCMC calibration outputs.

### Asymptomatic infections

We estimated that 74% of infections proceeded asymptomatically (70-78%, 95% PI) (see **Figure 2A**). Our model estimated that in total 1,304 (1,198-1,416) individuals were infected, representing 35% (32–38%) of the initial total population on the DP. Over half of these infections had not been detected at disembarkation on February 21st (53% (51-56%) **Figure 2C**), consisting of infected individuals who had recovered and became test negative before they were tested (37%, 34-40%), were yet to be tested (15%, 13-16%), or had recently been exposed and were not yet detectable at that point (1%, 1-3%). Nearly two-thirds of pre- and asymptomatic infections (67%, 66-68%) and 8% (6-9%) of symptomatic infections went undetected up until disembarkation (**Figure 2C**).

**Figure 2:**
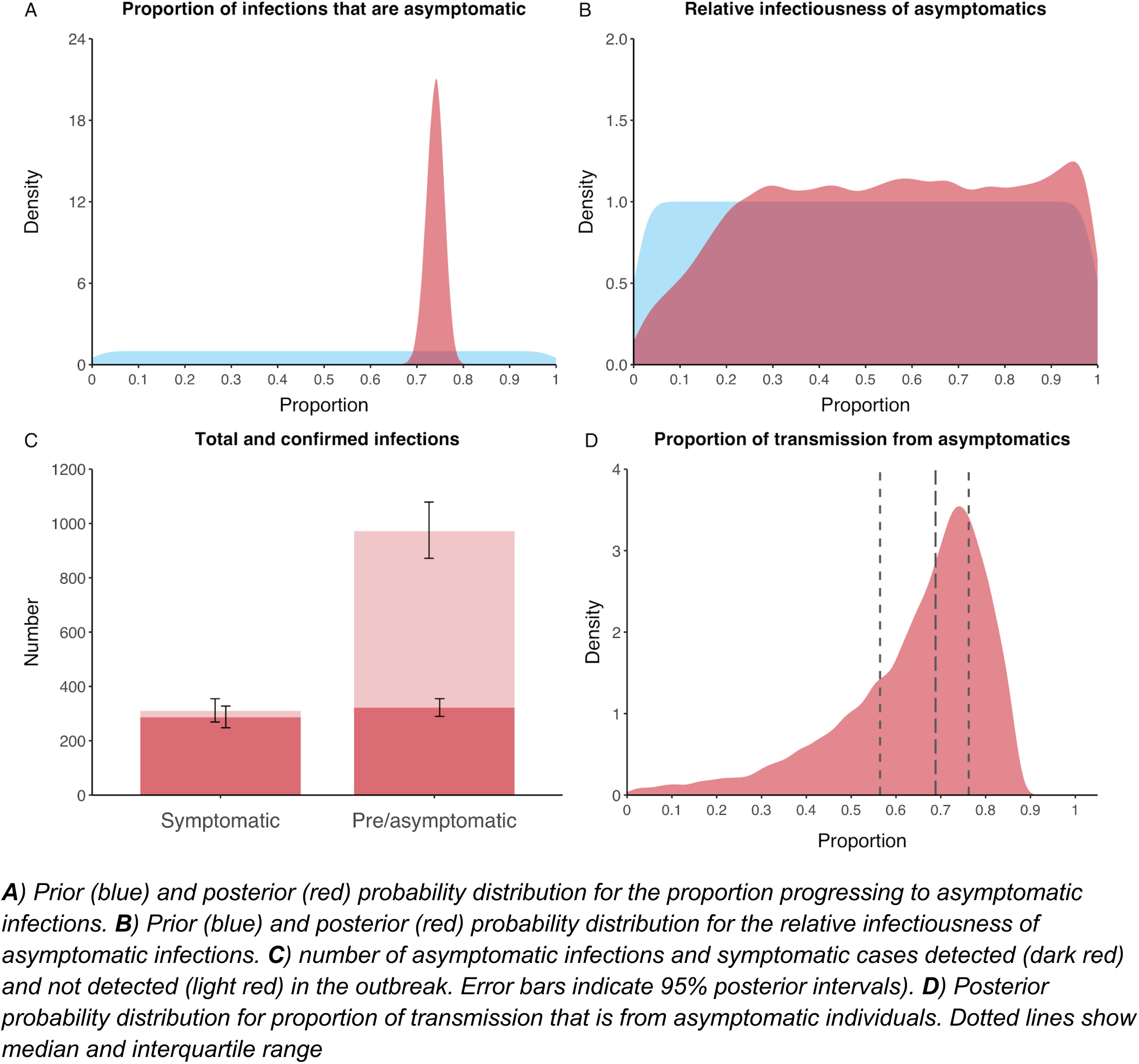
Proportion of asymptomatic infections, and contribution to transmission.

The model was unable to identify the relative infectiousness of asymptomatic infections from the data, i.e. a uniform prior was effectively returned (see **Figure 2B**). Combined with the estimated number of asymptomatic infections and the non-linear relationship between relative infectiousness and contribution to transmission (see supplementary materials, figure S1), the estimated proportion of transmission due to asymptomatic infections of 69% has a wide confidence interval (20-85%), although the IQR is 56%-76% (**Figure 2D**). The relative infectiousness of presymptomatics was also not identifiable, however, in all scenarios, the remaining transmission was equally distributed between the presymptomatic (14%, 1-44%) and symptomatic (17%, 11-42%) individuals.

Because of the non-identifiability of the relative infectiousness of asymptomatic infections we investigated marginal posterior estimates (Table 2). We find that low relative infectiousness of asymptomatic infections (0-25% compared to symptomatic individuals) would need to be compensated by a net reproduction number for individuals during their presymptomatic and symptomatic phase of 15.5 - 29.1.

**Table 2:** Model outputs by relative infectiousness of asymptomatic individuals.

| Range of relative infectiousness of asymptomatic individual | Model output |  |  |
| --- | --- | --- | --- |
|  | Transmission from asymptomatic individuals (%) | Net reproduction number for presymptomatic passengers | Basic reproduction number |
| 0-1% | 0-3 | 22.7 - 29.1 | 6.7 - 7.6 |
| 1-25% | 7-58 | 15.5 - 25.5 | 7.0 - 8.8 |
| 25-50% | 44-75 | 11.1 - 17.6 | 8.0 - 9.6 |
| 50-75% | 60-82 | 8.7 - 13.6 | 8.7 - 10.2 |
| 75-99% | 68-86 | 7.2 - 11.4 | 9.3 - 10.8 |
| 99-100% | 72-87 | 6.7 - 10.2 | 9.5 - 10.9 |

*Relative infectiousness expressed as proportion compared to symptomatic individuals. All values 95% posterior ranges from model scenarios. Net reproduction number represents the typical number of infections generated by a single individual during the presymptomatic and symptomatic stages.*

### Sensitivity analyses

Without an asymptomatic state the model was unable to reconstruct the dynamics of the outbreak (**Supplementary Figure 5**, Deviance Information Criterion (DIC) = 974 vs 329 for primary analysis). Adjusting the relative value for mixing between crew and passengers did not have a qualitative effect on the results (**Supplementary Figures 9 and 13)**.

When we assumed a fixed age-specific ratio for the proportion of infections that progress asymptomatically, the model was able to fit the data, although the number of correlated parameters was high. Overall results were similar to the main analysis, with a proportion asymptomatic of 42% (41-44%) and 89% (85-91%) for passengers and crew, respectively. The proportion of all transmission from asymptomatics was 69% (IQR = 59-74%). Relative infectiousness was again unidentifiable. See **Supplementary Figure 28** for details.

A longer latent period provided a poorer fit to the data (DIC = 361) (Supplementary Figure 24) Adjusting the duration of the asymptomatic state to half or double the sum of the presymptomatic and symptomatic states made little qualitative difference to the results (Supplementary Figures 17 and 21), although the shorter asymptomatic period was a marginally poorer fit to the data (DIC = 338). Finally, recalibrating the model assuming the 35 confirmed pre/asymptomatic cases where a test date was not available were allocated to the last feasible day (13th Feb) made no qualitative difference to our results (see **Supplementary Figure 33**).

## Discussion

### Summary

We find that in this well-documented outbreak in a closed population, 74% (70-78%) of infections proceeded asymptomatically, equaling a 1:3.8 (1:3.3-1:4.4) case-to-infection ratio. The majority of asymptomatic infections remained undetected, but may have contributed substantially to ongoing transmission. While the relative infectiousness of asymptomatic infections could not be identified, low infectiousness (e.g. 0-25% compared to symptomatic individuals) would have required a very high net reproduction number for individuals during their presymptomatic and symptomatic stages of (15.5 - 29.1).

### Interpretation

Our results are strongly informed by data, which show that when extensive symptom-agnostic testing was ramped up, substantial numbers of pre- or asymptomatic infections were identified. Given the clear suppression of transmission through quarantining, as indicated by the drop in incident symptomatic disease, this finding is most likely explained by a large proportion of undetected asymptomatic individuals.

The model and data were unable to identify a value for the relative infectiousness, although we showed how different ranges for this key parameter required specific trade-offs, as reflected in the basic reproduction number for infected individuals who will develop symptomatic disease. One can argue that a net reproduction number for presymptomatic passengers at the start of the outbreak of over 20 in this population, as required if asymptomatic individuals are effectively unable to transmit (range for relative infectiousness of 0-1%) is unlikely. Such high reproductive numbers are not usually seen, exceeding for example values found for norovirus outbreaks on cruise ships. (30) cruise While SARS-CoV-2 has been shown to survive on surfaces, (31) but this does not seem to be the primary mode of transmission. In combination with growing evidence around viral load in asymptomatic infections and their involvement in transmission chains (14), anecdotal evidence about transmission from asymptomatic individuals (19,32) including in closed populations, (33) it is reasonable to assume that asymptomatic infections play some role in ongoing SARS-CoV transmission. In our model, 83% of scenarios compatible with the data, asymptomatic infections were responsible for more than half of all transmission.

### Comparison to other studies

Our estimated proportion of asymptomatic infections in this outbreak is higher than previous studies, which relied on diagnosed cases only. (13) As we have shown, a substantial number of infections were not detected, which would explain some of the difference. Other empirical studies have found usually lower values, while some found similar ranges. While underestimation in other estimates due to low (17) and imbalanced participation from individuals with and without symptoms (18) will be part of the explanation, there remains scope for unexplained variation from more complete samples. (14) In addition, it is possible that PCR-based testing has a lower sensitivity for asymptomatic infections, which would further increase the proportion of asymptomatic infections. (32)

A sensitivity analysis showed that our results were robust to age-specific probabilities of progressing to asymptomatic infections, as well as other assumptions made by the model, and driven by trends in the data.

Our estimated substantial contribution to transmission from asymptomatic infections confirms an hypothesis from Nishiura after analysing symptomatic cases occurring post-disembarkation. (21) Our initial reproduction number of 9.3 (7.4-10.6) reflects the high transmission environment on cruise ships, although lower that found in an earlier analysis by Rocklöv et al. (34)

Our finding of similar contribution to transmission from presymptomatic and symptomatic individuals also matches findings by others. (5,11,12) In line with this, it is clear that having symptoms, or at least being aware of them, is not required in the transmission of SARS-CoV-2. (5,11,12,35,36) Although cough is often considered essential for transmission of respiratory infections (37), work in tuberculosis, influenza and other coronaviruses has shown that while a cough may increase spread, it is not a requirement. Transmission from breathing, talking and sneezing is also possible, as well as transmission from contaminated surfaces. (31,38–40)

### Limitations

Additional data, in particular on the distribution of asymptomatic infections across crew and passengers, by age and shared quarantine environments would have benefited the model and potentially enable us to estimate a range for the relative infectiousness of asymptomatic infections. Also, a serological survey of the population, and the date and testing history of individuals who developed symptoms post-disembarkation would have likely informed more precise model estimates. In addition, better evidence on performance of the test used, and the associated likelihood of false-negative or false-positive results would help refine estimates. As more data becomes available, future model analyses of SARS-CoV dynamics in closed populations should further inform the key questions we have looked to address here.

## Conclusion

Asymptomatic SARS-CoV-2 infections may contribute substantially to transmission. This is essential to consider for countries when assessing the potential effectiveness of ongoing control measures to contain COVID-19.

## Data Availability

All data and model code are publicly available. See supplementary materials, and github repository in the link provided

https://cmmid.github.io/topics/covid19/asymp-transmission.html

## Acknowledgements

The authors would like to thank Dr Taichi Kidani for help in translating primary data sources.

The following funding sources are acknowledged as providing funding for the named authors. This research was partly funded by the Bill & Melinda Gates Foundation (INV-003174: YL; NTD Modelling Consortium OPP1184344: CABP). DFID/Wellcome Trust (Epidemic Preparedness Coronavirus research programme 221303/Z/20/Z: CABP). ERC Starting Grant (#757699: JCE, RMGJH). This project has received funding from the European Union’s Horizon 2020 research and innovation programme - project EpiPose (101003688: YL). HDR UK (MR/S003975/1: RME). This research was partly funded by the National Institute for Health Research (NIHR) using UK aid from the UK Government to support global health research. The views expressed in this publication are those of the author(s) and not necessarily those of the NIHR or the UK Department of Health and Social Care (16/137/109: YL). UK MRC (MC_PC 19065: RME; MR/P014658/1: GMK). Wellcome Trust (206250/Z/17/Z: AJK, TWR; 208812/Z/17/Z: SFlasche; 210758/Z/18/Z: JH, SFunk).

The following funding sources are acknowledged as providing funding for the CMMID 2019 nCoV working group. Alan Turing Institute (AE). BBSRC LIDP (BB/M009513/1: DS). This research was partly funded by the Bill & Melinda Gates Foundation (INV-003174: KP, MJ; NTD Modelling Consortium OPP1184344: GM; OPP1180644: SRP; OPP1183986: ESN; OPP1191821: KO’R, MA). DFID/Wellcome Trust (Epidemic Preparedness Coronavirus research programme 221303/Z/20/Z: KvZ). Elrha R2HC/UK DFID/Wellcome Trust/This research was partly funded by the National Institute for Health Research (NIHR) using UK aid from the UK Government to support global health research. The views expressed in this publication are those of the author(s) and not necessarily those of the NIHR or the UK Department of Health and Social Care (KvZ). ERC Starting Grant (#757688: CJVA, KEA; #757699: MQ). This project has received funding from the European Union’s Horizon 2020 research and innovation programme - project EpiPose (101003688: KP, MJ, PK, WJE). This research was partly funded by the Global Challenges Research Fund (GCRF) project ‘RECAP’ managed through RCUK and ESRC (ES/P010873/1: AG, CIJ, TJ). Nakajima Foundation (AE). NIHR (16/137/109: BJQ, CD, FYS, MJ; Health Protection Research Unit for Modelling Methodology HPRU-2012-10096: NGD, TJ; PR-OD-1017-20002: AR). Royal Society (Dorothy Hodgkin Fellowship: RL; RP\EA\180004: PK). UK DHSC/UK Aid/NIHR (ITCRZ 03010: HPG). UK MRC (LID DTP MR/N013638/1: EMR, QJL). Authors of this research receive funding from UK Public Health Rapid Support Team funded by the United Kingdom Department of Health and Social Care (TJ). Wellcome Trust (208812/Z/17/Z: SC; 210758/Z/18/Z: JDM, NIB, SA, SRM). No funding (AKD, AMF, DCT, SH).

## Author Contributions

RMGJH conceived the study. JCE, SF and RMGJH designed the model with TWR, YL, JH, CABP providing input. JCE processed the data, designed the software and inference framework and implemented the model. RMGJH and JCE wrote the first draft of the manuscript. All authors interpreted the results, contributed to writing, and approved the final version for submission.

## Notes

### Competing Interest Statement

The authors have declared no competing interest.

### Funding Statement

Funding: ERC Starting Grant (#757699), Wellcome trust (208812/Z/17/Z), HDR UK (MR/S003975/1)
The funder of the study had no role in study design, data collection, data analysis, data interpretation, or writing of the report. The corresponding author had full access to all the data in the study and had final responsibility for the decision to submit for publication.

